# Diagnostic value of combined detection of pepsinogen, gastrin-17, and ^13^C-urea breath test in children with chronic gastritis: A multivariate analysis

**DOI:** 10.1101/2025.08.18.25333943

**Authors:** Yiyun Gao, Chuangui Liu, Kaishi Oayang, Siqi Li, Weilin Huang

**Affiliations:** Department of Clinical Laboratory, Jun’an Branch Hospital of Shunde Hospital of Guangzhou University of Chinese Medicine, No. 7 Bai’an Road, Foshan, Guangdong 528329, China; Department of Dermatology, Jun’an Branch Hospital of Shunde Hospital of Guangzhou University of Chinese Medicine, No. 7 Bai’an Road, Foshan, Guangdong 528329, China; Department of Anesthesiology, Shunde Hospital of Guangzhou University of Chinese Medicine, No.12 Jinsha Avenue, Foshan, Guangdong 528300, China

**Keywords:** Gastrin-17, Pepsinogen, ^13^C-urea breath test, Chronic gastritis in children, Multivariate logistic regression analysis

## Abstract

**Background:** This study evaluated the diagnostic efficacy of combining pepsinogen (PG I/II), gastrin-17 (G-17), and ^13^C-urea breath test (^13^C-UBT) for chronic gastritis in children using logistic regression and receiver operating characteristic (ROC) analysis.

**Methods:** Between June 2018 and August 2023, 65 children from Jun’an Branch Hospital of Shunde Hospital of Guangzhou University of Chinese Medicine, diagnosed with chronic gastritis (chronic gastritis group), and 50 healthy children (control group) participated in this study. The enzyme-linked immunosorbent assay (ELISA) determined serum levels of PG I, PG II, and G-17. Furthermore, ^13^C-UBT was performed for *Helicobacter pylori* (Hp) infection detection. Both groups underwent serological tests and gastroscopy.

**Results:** Combined detection showed significantly higher positive rates than individual PG I/II or G-17 tests (P < 0.05), though comparable to ^13^C-UBT alone. Chronic gastritis patients exhibited elevated PG I, PG II, and G-17 levels versus controls (P < 0.05). Multivariate analysis identified PG I (OR = 2.982, P = 0.011), G-17 (OR = 3.527, P = 0.0013), and ^13^C-UBT positivity (OR = 4.193, P = 0.002) as significant predictors. ROC analysis revealed AUCs of 0.673 (PG I), 0.792 (G-17), 0.814 (^13^C-UBT), and 0.887 (combined), with sensitivity ≥89% and specificity >76%.

**Conclusion:** Serum PG I, G-17 detection, and ^13^C-UBT provided considerable predictive accuracy for chronic gastritis in children, and combination testing further improved diagnostic accuracy in this population.

## 1. Introduction

Chronic gastritis (CG) is a prevalent digestive system disorder characterized by prolonged inflammation of the gastric mucosa. It is primarily classified into superficial, atrophic, and specific forms of gastritis [1]. The pathological manifestations include damage to the gastric mucosal epithelium, a reduction in glandular structures, and infiltration of inflammatory cells. Chronic gastritis arises from multiple factors, including *Helicobacter pylori* (Hp) infection, lifestyle choices (e.g., smoking, alcohol use), dietary patterns, and nonsteroidal anti-inflammatory drugs (NSAIDs) overuse [2, 3]. Epidemiological data reveal a significant global prevalence of chronic gastritis, especially in middle-aged and elderly populations, with incidence rates rising with age. The clinical symptoms typically include epigastric discomfort, nausea, and loss of appetite, though some patients may remain asymptomatic. In recent years, the incidence of chronic gastritis in children has also risen, with regional incidence rates in China ranging between 45% and 80% due to variations in dietary patterns [4, 5]. The gastric mucosa of children is comparatively fragile, making it more susceptible to damage and increasing the risk of gastritis. Early prevention and therapy in pediatric populations are essential because they can reduce the likelihood that serious gastrointestinal diseases will develop in adulthood. Gastroscopy offers clinicians direct diagnostic evidence for chronic gastritis; however, challenges such as incomplete physiological development, low compliance, and safety concerns related to anesthesia restrict its application in pediatric populations [6, 7]. Therefore, developing more practical diagnostic methods for chronic gastritis in pediatric patients is essential. Serological markers, including pepsinogen I (PG I) and pepsinogen II (PG II), indicate the condition of intestinal mucosa. A diminished PG I/PG II ratio (PGR) generally signifies atrophic gastritis, a disease intimately associated with later stomach carcinogenesis [8]. Gastrin-17 (G-17), a precursor of gastrin, may signal disease progression of chronic gastritis toward pre-cancerous lesions when elevated [9]. Furthermore, the non-invasive ^13^C-urea breath test (^13^C-UBT) holds significant value in diagnosing and managing Hp infection, a key contributor to chronic gastritis [10].

This study includes pediatric patients diagnosed with chronic gastritis from the gastroenterology department, alongside healthy children serving as controls. The analysis of PG I/II, G-17, and ^13^C-UBT results using various mathematical-statistical models aimed to assess the clinical utility of these established serological and breath-test markers for the screening and diagnosing of chronic gastritis in children.

## 2. Materials and Methods

### 2.1. General information

A total of 65 children (32 boys, 33 girls; mean age 8.9 ± 2.4 years) diagnosed with chronic gastritis at Jun’an Branch Hospital of Shunde Hospital of Guangzhou University of Chinese Medicine between 23 June 2018 and 29 August 2023 were enrolled. The sample size was determined based on comparable diagnostic studies [11].

#### Inclusion criteria for the chronic gastritis group required

1. Endoscopic and histopathological confirmation of chronic gastritis;
2. Age 6-12 years.

#### Exclusion criteria comprised

1. Severe cardiopulmonary dysfunction or major systemic diseases;
2. History of gastroduodenal surgery;
3. Use of proton pump inhibitors (PPIs)/antibiotics within 1 month or gastroprotective agents within 1 week;
4. Prior *H. pylori* eradication therapy;
5. Any condition contraindicating participation.

For comparative analysis, 50 asymptomatic controls (24 boys, 26 girls; mean age 9.2 ± 2.1 years) were recruited as control group.

#### Control group inclusion mandated

1. Age 6-12 years.
2. Absence of gastrointestinal symptoms (abdominal pain, nausea, dyspepsia) for ≥6 months;
3. Normal physical examination and laboratory parameters (complete blood count, hepatic/renal function);
4. Macroscopically normal gastric mucosa confirmed by painless gastroscopy without biopsy (Section 2.2.3).

#### Identical exclusion criteria were applied to both cohorts

### 2.2 Methods

#### 2.2.1. Serological test

Serological tests were performed in both groups. Subjects were prevented from food intake after 20:00 the evening before blood collection. The following morning, 5 mL of peripheral blood was collected from the median cubital vein and placed into vacuum tubes. Samples underwent centrifugation at 3000 rpm for 10 min to isolate serum, which was stored at −80°C. Commercial ELISA kits (Biohit Oyj, Finland for pepsinogen; Zhejiang ERKN Biotechnology, China for gastrin) were utilized to quantify serum PG I, PG II, and G-17 levels.

#### 2.2.2. ^13^C-urea breath test

After fasting blood collection, baseline breath samples were obtained. Subjects then ingested a ^13^C -urea capsule (Beijing Boran Pharmaceutical Co., Ltd.) with water. Post-dose breath samples were collected 30 min later. The two samples were analyzed using the HY-IREXA ^13^C breath analyzer (Guangzhou Huayou Mingkang Optoelectronics Co., Ltd.).

#### 2.2.3. Gastroscopy

Gastroscopies were performed in both groups. Control group participants underwent gastroscopy to confirm the absence of gastric pathology. Subjects commenced fasting at 20:00 the evening before the examination. After overnight fasting, painless gastroscopy was performed using an Olympus CV-290 endoscope (Japan). Subjects were anesthetized with intravenous propofol (2 mg/kg). If inflammatory lesions were detected during gastroscopy, 3–5 biopsy samples were collected from the gastric antrum, body, and lesion sites. The collected biopsy samples were immobilized with 37%–40% formaldehyde solution and sent to the pathology department for pathological examination. The biopsy samples were paraffin-embedded, sectioned, and stained with hematoxylin-eosin (H&E) for pathological assessment by certified pathologists. The definitive diagnosis was determined by histological findings.

### 2.3. Statistical Analysis

The data were statistically processed using SPSS 19.0 software. Quantitative values are expressed as Mean ± SEM. One-way ANOVA was employed to compare differences among different groups with data showing normal distribution and homogeneity of variance. Categorical data are displayed as frequency/percentage, and inter-group comparisons were conducted using the χ2 test. A logistic regression model was developed for multifactorial regression analysis. The predictive accuracy of each test item for chronic gastritis in children was calculated. Diagnostic performance of biomarkers (PG I/II, G-17, ^13^C-UBT) was evaluated via ROC curve analysis. A *P* value of < 0.05 was considered statistically significant. All quantitative variables (e.g., PG I, PG II, G-17 serum levels) were analyzed as continuous variables. No grouping or transformation was applied to these variables to preserve their original distribution and maximize statistical power.

## 3. Results

### 3.1 Demographic statistics and positive rate comparisons

This study included 65 children with chronic gastritis (32 boys, 33 girls) and 50 healthy controls (24 boys, 26 girls). All participants underwent a painless gastroscopy under intravenous propofol anesthesia along with three additional gastroenterological examinations. In the entire cohort, the positive rates for individual tests (PG I/II, G-17, ^13^C-UBT) and combined detection (defined as positivity in any of the three tests) were 12.17%, 15.65%, 30.43%, and 34.78%, respectively. The positive rate for combined detection was significantly higher than individual PG I/II (χ^2^ = 16.359, *P* < 0.01) and G-17 tests (χ^2^ = 11.159, *P* < 0.01) but comparable to ^13^C-UBT alone (χ^2^ = 0.49462, *P* = 0.4819).

### 3.2. Gastroscopic and pathological findings

Painless gastroscopy demonstrated normal results in all 50 healthy controls. On the other hand, all 65 children in the chronic gastritis group were diagnosed with chronic gastritis through gastroscopic and histopathological examinations.

### 3.3. Comparison of positive rates between individual tests and combined detection

Compared to the control group, the chronic gastritis group showed significantly higher positive rates for individual test of G-17 and ^13^C-UBT, and combined detection (*P* < 0.05). The PG I/II individual test in the chronic gastritis group showed a higher positive rate than in the control group but not statistically significant (Table 1).

**Table 1.** Comparison of Positive Rates for individual PG I/II, G-17, ^13^C-UBT assays, and combined detection in pediatric chronic gastritis [n (%)].

| Group | Number of Cases | Positive Rate for PG I/II Test | Positive Rate for G-17 Test | Positive Rate for $^{13}\text{C}$-UBT | Positive Rate for Combined Detection |
| --- | --- | --- | --- | --- | --- |
| Control Group | 50 | 4 (8.00) | 3 (6.00) | 10 (20.00) | 11 (22.00) |
| Chronic Gastritis Group | 65 | 10 (15.38) | 15 (23.08) | 25 (38.46) | 29 (44.62) |
| $\chi^2$ | | 1.44 | 6.24 | 4.55 | 6.37 |
| $P$ | | 0.2299 | 0.0125 | 0.0329 | 0.0116 |

### 3.4. Comparison of serum PG I/II and G-17 levels between groups

PG I and PG II levels in chronic gastritis group were significantly higher than those in control group (*P* < 0.05). The calculated PG I/PG II ratio (PGR) was lower in the chronic gastritis group, but the difference lacked statistical significance (Table 2).

**Table 2.** Comparison of PG I, PG II, PGR, and G-17 levels between groups [Mean ± SEM].

| Group | Number<br>of cases | PG I ($\mu\text{g/L}$) | PG II ($\mu\text{g/L}$) | PGR | G-17 (pmol/L) |
| --- | --- | --- | --- | --- | --- |
| Control<br>Group | 50 | 106.38±18.14 | 13.73±6.03 | 8.04±1.98 | 5.34±1.21 |
| Chronic<br>Gastritis<br>Group | 65 | 121.45±15.04* | 18.26±5.18* | 7.94±2.03 | 9.93±1.78** |
| $\chi^2$ | | | | | |
| *P* |  | < 0.05 | < 0.05 | > 0.05 | < 0.05 |

\*Note: Compared with control, \* $P$ <0.05, \*\* $P$ <0.01.

### 3.5. Multivariate logistic regression analysis of PG I/II, G-17, and 13C-UBT in predicting chronic gastritis in children

According to the results of multivariate logistic regression analysis, positive levels of PG I, PG II, G-17, and ^13^C-UBT were all potential indicators for predicting chronic gastritis in children. PG-I substantially impacted the disease, as evidenced by its regression coefficient of 1.012 and a significant *P* value of 0.011 (OR = 2.982, 95% CI: 0.734-4.262). Although PG II had a comparatively high regression coefficient (1.372), there was no statistical significance shown by its *P* value of 0.126. With a substantial impact on the disease (OR = 3.527, 95% CI: 0.419-6.732), G-17 had a P value of 0.0013. ^13^C-UBT positivity (OR = 4.193, *P* = 0.002) was also a significant predictor of chronic gastritis (Table 3).

**Table 3.** Multivariate logistic regression analysis of PG I/II, G-17, and ^13^C-UBT for pediatric chronic gastritis.

| Indicator | Regression Coefficient | Standard Error | z-value | Wald $\chi^2$ | P-value | OR-value | 95% CI |
| --- | --- | --- | --- | --- | --- | --- | --- |
| PG I | 1.012 | 0.336 | 1.374 | 3.219 | 0.011 | 2.982 | 0.734 - 4.262 |
| PG II | 1.372 | 0.355 | 1.212 | 2.058 | 0.126 | 1.512 | 0.442-3.837 |
| G-17 | 1.223 | 0.673 | 1.672 | 3.345 | 0.0013 | 3.527 | 0.419-6.732 |
| <sup>13</sup>C-UBT Positive | 0.973 | 0.234 | 0.794 | 0.683 | 0.002 | 4.193 | 0.634-5.339 |

### 3.6 Diagnostic value of PG I/II, G-17, and ^13^C-UBT indicators

ROC analysis revealed that the area under the curve (AUC) for PG II, PG I, G-17, and ^13^C-UBT was 0.579 (95%CI: 0.364∼0.662, *P* > 0.05), 0.673 (95%CI: 0.356∼0.734, P < 0.05), 0.79 (95%CI: 20.521∼0.842, P < 0.05), and 0.814 (95%CI: 0.415∼0.833, P < 0.05), respectively (Figure 1). The combined detection (positivity defined as PG I/II/G-17 exceeding optimal thresholds and 13C-UBT positivity) outperformed the individual tests with an AUC of 0.887, a sensitivity of 89%, and a specificity of 76%. (Table 4).

**Table 4.**
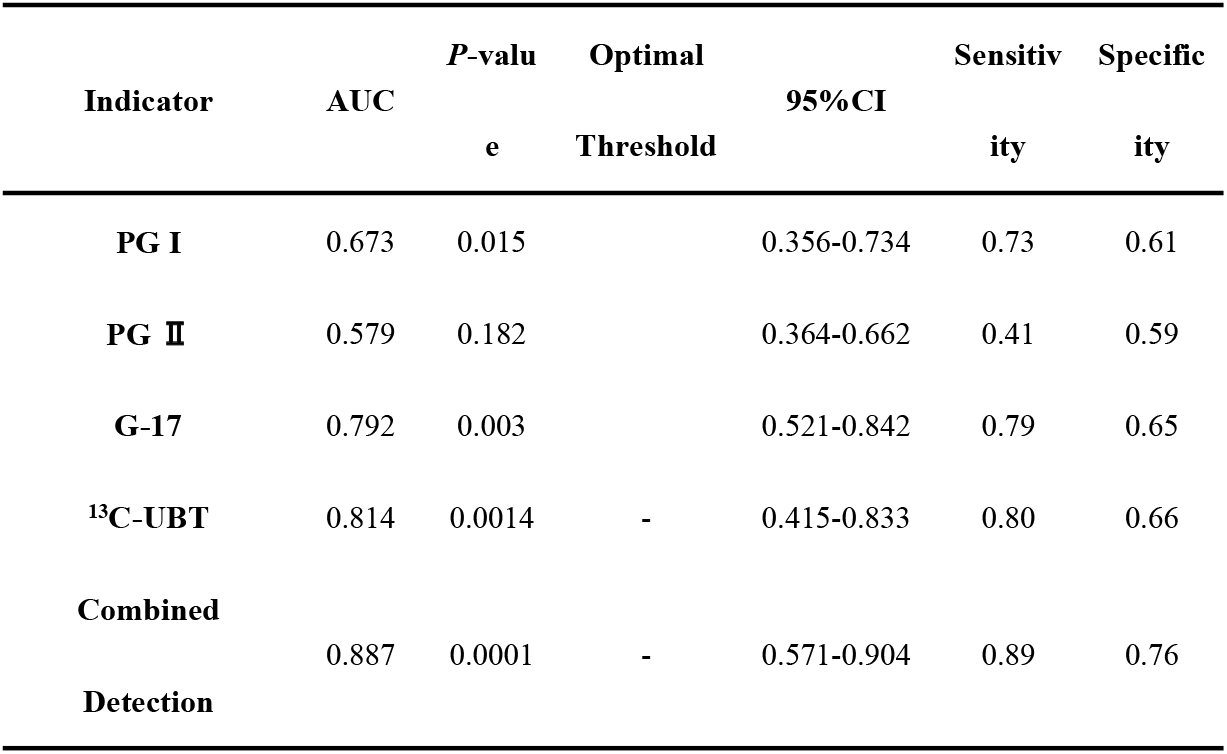
AUC Diagnostic Value of PG I/II, G-17, ^13^C-UBT, and Combined Detection.

| Indicator | AUC | *P*-value | Optimal Threshold | 95%CI | Sensitivity | Specificity |
| --- | --- | --- | --- | --- | --- | --- |
| **PG I** | 0.673 | 0.015 |  | 0.356-0.734 | 0.73 | 0.61 |
| **PG II** | 0.579 | 0.182 |  | 0.364-0.662 | 0.41 | 0.59 |
| **G-17** | 0.792 | 0.003 |  | 0.521-0.842 | 0.79 | 0.65 |
| **<sup>13</sup>C-UBT** | 0.814 | 0.0014 | - | 0.415-0.833 | 0.80 | 0.66 |
| **Combined Detection** | 0.887 | 0.0001 | - | 0.571-0.904 | 0.89 | 0.76 |

**Figure 1.**
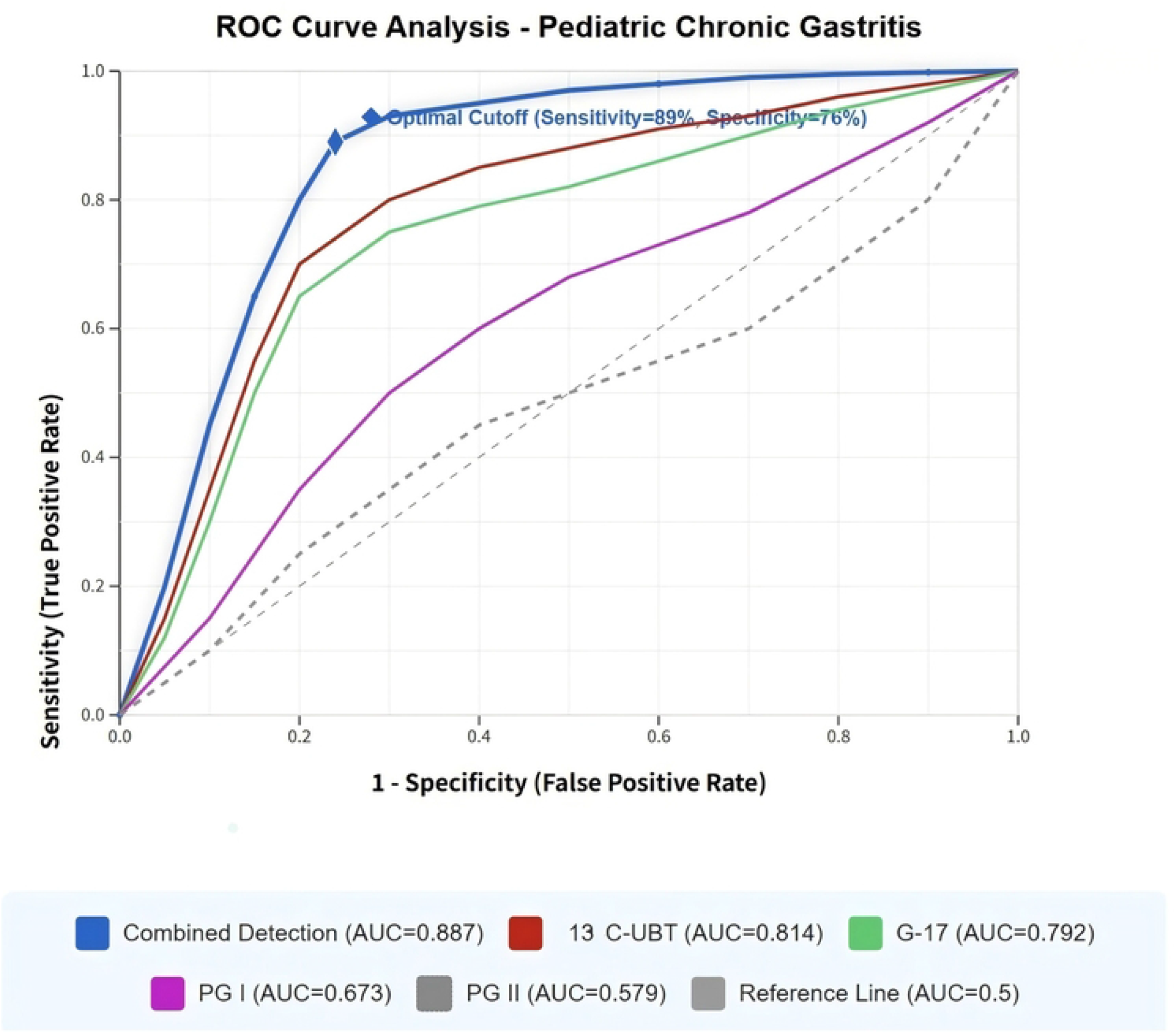
ROC Curve Analysis.

## 4. Discussion

The causes of chronic gastritis in children are different from those in adults. Hp infection is a primary causative factor due to their immature immune systems, though dietary habits, environmental influences, and genetic susceptibility also play significant roles [12]. The fragile gastric mucosa in children enhances susceptibility to extended inflammation, potentially disrupting gastric acid secretion and resulting in dyspepsia and impaired nutrient absorption. Persistent damage to gastric mucosa may lead to atrophy, elevating the risk of gastric ulcers and other severe gastrointestinal disorders. Chronic gastritis negatively affects the quality of life in children, presenting as recurrent abdominal pain, anorexia, and failure to thrive [13, 14]. Early diagnosis and intervention are thus critical to prevent disease progression and long-term sequelae.

Our findings demonstrate that the multi-marker panel (PG I/II + G-17 + ^13^C-UBT) achieved an AUC of 0.887, outperforming individual tests (AUC range: 0.579–0.814), suggesting synergistic diagnostic value. The elevated positive rate of combined testing in the chronic gastritis group, particularly its alignment with ^13^C-UBT outcomes, highlights the critical importance of Hp in the pathogenesis of pediatric gastritisWhile individual biomarkers hold diagnostic value, their synergistic use enhances accuracy and reduces missed diagnoses.

In agreement with previous studies, PG I and PG II levels demonstrate a strong correlation with gastric mucosal dysfunction. Existing literature highlights their utility in monitoring mucosal atrophy, especially during early-stage carcinogenesis [15]. Furthermore, G-17, an essential indicator of gastric acid secretion, shows significant variation in individuals with gastric acid secretion disorders. Research indicates that G-17 may possess substantial diagnostic value in chronic gastritis, particularly in pediatric patients with acid-related abnormalities [11].

Hp infection is one of the important causes of chronic gastritis. Although not all infected individuals develop gastritis, most chronic Hp infections lead to persistent inflammation of the stomach. The ^13^C-UBT, extensively used in diagnosing gastrointestinal disorders, validated the association between Hp and chronic gastritis. Routine screening of Hp infection can effectively prevent and cure gastritis and its complications [13]. Moreover, combined biomarker assessment provides a holistic evaluation of mucosal health, facilitating early diagnosis [16].

This study validated the diagnostic value of the combined assessment of the three indications. However, certain limitations exist. The limited size and singular focus of the cohort restrict the generalizability of the findings. Results may require multicenter validation due to regional dietary variations. Future research should expand sample sizes and incorporate diverse locations and demographics to improve representativeness. Additionally, the high false-positive rate observed in the control group for ^13^C-UBT, and combined testing may be attributed to:(1) Transient mucosal inflammation induced by dietary or medication stimuli in the control group [17]; (2) Asymptomatic *H. pylori* colonization without histological manifestations of gastritis [18]. Furthermore, due to the distinct pathogenesis of gastritis in children compared to adults, particularly regarding the presentation of Hp infection and the reparative capacity of gastric mucosa, it is crucial to investigate the pathophysiological variations among children of varying ages.

Clinically, early detection of chronic gastritis is critical to prevent irreversible mucosal damage. Non-invasive approaches, such as combined assessment of PG I/II, G-17, and ^13^C-UBT, provide a streamlined diagnostic modality for pediatric gastritis. These serological and breath-based assays demonstrate significantly lower invasiveness than conventional gastroscopy and are particularly suitable for population-level screening in pediatric cohorts. Future research should focus on: (1) validating their utility in monitoring therapeutic responses, and (2) elucidating associations with comorbid gastrointestinal disorders (e.g., peptic ulcer disease, functional dyspepsia). Advances in precision medicine incorporating molecular and genetic biomarkers may further refine diagnostic and therapeutic algorithms. Additionally, screening for virulent *Helicobacter pylori* strains (e.g., Cag A+) in infected children should be prioritized given their established association with gastric carcinogenesis.

## 5. Conclusion

In conclusion, the combined detection of PG I/II, G-17, and ^13^C-UBT demonstrates promising diagnostic potential for chronic gastritis in children. While our findings provide robust evidence, broader validation of its clinical applicability and long-term benefits is warranted. Continued innovation in non-invasive technologies will further enhance the convenience and efficacy of early screening and intervention for chronic gastritis in children.

## Data Availability

All data in the current study are available from the corresponding author on reasonable request.

## Acknowledgements

None.

## Abbreviations

^13^C-UBT: ^13^C-urea breath test
ROC: Receiver operating characteristic
ELISA: Enzyme-linked immunosorbent assay
Hp: *Helicobacter pylori*
CG: Chronic gastritis
NSAIDs: Nonsteroidal anti-inflammatory drugs
PG I: Pepsinogen I
G-17: Gastrin-17
PPIs: Proton pump inhibitors

## Ethics approval and consent to participate

This retrospective study was approved by the Ethics Committee of Jun’an Branch Hospital of Shunde Hospital of Guangzhou University of Chinese Medicine (Approval No. KT-202306) with waived informed consent, complying with national regulations for retrospective research using fully anonymized data. The authors did not have access to identifiable participant information during or after data collection and the ethics committee confirmed that no direct subject contact or identifiable information access occurred during this analysis. Study procedures adhered to the Declaration of Helsinki and Chinese ethical guidelines. This study was registered with the Chinese Clinical Trial Registry (Registration No. MR-44-24-003621).

## CRediT authorship contribution statement

**Yiyun Gao**: Writing - original draft, Methodology, Conceptualization. **Chuangui Liu**: Formal analysis. **Kaishi Oayang**: Writing - original draft. **Siqi Li**: Formal analysis. **Weilin Huang**: Writing – review & editing, Validation, Supervision.

